# Clinical Evaluation of Post-Surgical Scar Hyperesthesia; an Exploratory Longitudinal Study

**DOI:** 10.1101/2023.03.25.23287735

**Authors:** Donna L. Kennedy, Shehan Hettiaratchy, Caroline M. Alexander

## Abstract

Evidence for the objective clinical evaluation of scar hyperesthesia is lacking. This exploratory study investigated the clinical relevance and responsiveness of objective scar evaluation measures in adults following hand surgery.

With ethical approval and consent, participants were enrolled from one NHS hospital. Patient reported and investigator completed scar morphology, cosmesis, pain and function were evaluated at 1- and 4-months post-surgery. Statistical analysis investigated the responsiveness of outcome measures and association of physical measures with the Palmar Pain Severity Scale (PPS).

21 participants enrolled prior to premature study closure due to the COVID-19 pandemic; 13 completed follow up. Scar pain (p=.002); scar interference (PPI [p=.009]) and Brief Pain Inventory (BPI) scores (p=.03) improved. Neuropathic Pain Symptom Inventory (NPSI) scores demonstrated heterogeneity in scar pain; evoked pain predominated. Patient Scar Assessment Questionnaire (PSAQ) indicated improvement in cosmetic dissatisfaction and consciousness (p=.03; p=.003), respectively. Baseline psychological screening scores correlated with scar pain (p=.04), and interference (p< .001). Scar morphology, pliability and inflammation were not associated with scar pain. Significant differences in scar mechanical pain sensitivity (p=.04) and cold pain threshold (p=.05) were identified.

PPS and PPI scores were responsive in a heterogeneous hand surgery sample. BPI ‘worst pain’ identified severe pain, suggesting composite scar pain scores are required. The PSAQ robustly measured scar appearance and consciousness. Psychophysical tests of mechanical and thermal sensitivity are potential candidate objective measures of scar hyperesthesia. The NPSI demonstrates clinical utility for exploring scar pain symptoms and may support the elucidation of the drivers of persistent scar pain.

## BACKGROUND

When skin is wounded by injury or surgery, a scar results. It is estimated that over 100 million people develop scars yearly, primarily as a result of surgical procedures (1). Scarring may be burdensome and deleterious, causing unpleasant or painful symptoms which interfere with activities of daily living and social participation (2). The imperative to improve scar outcomes has resulted in the development of a vast array of scar treatments with variable efficacy (3-5); at present there is an estimated $12 billion market annually in the United States for scar treatment (6).

Scars are often persistently painful or hypersensitive, however, the evaluation of scar pain is not standardised (7-10) and currently there is no gold-standard outcome measure for evaluating scar (11). In practice, scar pain evaluation predominantly focuses on quantifying pain intensity with patient reported scales. Beyond intensity, evaluation generally fails to consider important pain parameters including symptoms, quality, and functional interference. The objective physical evaluation of scar pain has received little attention (12-14). Secondarily, the nature of scar pain and associated impairment is poorly understood.

Although the physical characteristics of scar are thought related to scar pain, there is no evidence for how the morphology of a hyperaesthetic scar differs from a quiescent scar (15, 16). Mechanisms underlying scar pain are not fully elucidated and are likely multifactorial. Scar pain may be related to the severity of tissue trauma and to psychological factors including anxiety (9). It is further theorised that scar pain is driven by an increase in small nerve fibre density; increased density of pro-inflammatory sensory neuropeptides promoting sensitisation; and mechanical compression of Aδ & C-fibres by dense scar tissue (9).

As an outcome, scar pain lacks a working definition, vital to consistency in assessment. The International Association for the Study of Pain (17) defines hyperesthesia as increased sensitivity to stimulation, including touch and thermal stimuli, that may or may not be painful (18). Utilising ‘scar hyperesthesia’ and it’s working definition in outcome evaluation may promote standardisation.

There is scant evidence for evaluating post-surgical scar hyperesthesia and this results in unquantified and unaddressed suffering, impairment, and participation restriction. This study aimed to investigate the nature of post-operative scar hyperesthesia and the clinical utility of objective physical scar outcome measures. Post-surgical (elective, or planned surgery) hand scars were chosen as a suitable model for investigation, as they are prevalent, relatively homogeneous, and physically accessible for examination. However, it is anticipated that study findings will be generalisable to surgical scars throughout the body.

### Study questions

▪ Is there variability in patient-reported scar pain symptoms, possibly indicating heterogeneity in underlying pathophysiological mechanisms?
▪ Are objective measures of thermal and mechanical pain threshold, scar inflammation and scar pliability associated with patient-reported scar pain?

### Study aims

To explore the nature of scar hyperesthesia and investigate the association of objective measures of scar morphology, somatosensory function, inflammation, and pliability with patient-reported scar hyperesthesia, thereby identifying robust, clinically relevant measures for future trials.

## METHODS

A longitudinal observational study conducted in accordance with the 18th World Medical Assembly, Helsinki 1964 and later revisions. Ethical approval was received from HRA and Health and Care Research Wales (HCRW) on 20^th^ December 2018 (18/LO/2161). Two patient-collaborators (MB; VG) reviewed study measures, procedures and documentation and provided feedback to improve rigour, transparency and clarity. National Health Service Trust pathways for patient care continued throughout. Sequential adult patients listed for elective hand surgery were recruited by poster, at clinic appointments and by post. Travel was reimbursed.

### Participation criteria

Adults over 18 years undergoing unilateral elective hand surgery were recruited. Exclusion criteria were inadequate English language to comprehend and consent to study measures, diagnosed serious medical or psychological comorbidity, surgery for a traumatic hand injury, post-surgical wound complication (i.e., infection), history of pathological scarring, previous hand or wrist trauma or surgery, neurological conditions, blood thinning medication, pregnancy – by patient report because fluctuating hormone levels may affect pain thresholds.

### Procedure

With informed, written consent, participants completed baseline assessment 4-weeks (± 14 days) post-surgery and follow-up 4-months (± 30 days) post-surgery. Four months post-surgery was identified as a relevant period, as tissue healing would be adequate for return to functional activity.

At baseline, demographic data and medical history were recorded. Outcome measures were repeated at baseline and follow-up, except pressure and thermal pain thresholds were assessed only at follow-up to avoid risk of injury in healing scar. At follow up, wound healing complications were recorded as well as information regarding clinician provided or participant-initiated scar care.

### Participant-completed questionnaires

Participant-completed questionnaires were used to evaluate scar outcome, pain parameters, and psychological comorbidity. Scar appearance, consciousness and satisfaction with symptoms were evaluated with the Patient Scar Assessment Questionnaire (PSAQ). (19) The PSAQ is a valid, reliable patient-completed outcome measure (10, 20, 21). Subscale items have 4-point categorical responses with 1 point for the most favourable category and 4 for the least favourable. The PSAQ appearance subscale has nine questions (score range 8-32), scar consciousness six questions (range 6-24) and satisfaction with symptoms five questions (score range 5-20). At follow-up, participants completed a 100 mm visual analogue scale of global scar outcome, as compared to the unaffected hand, with unaffected skin rated “100” and the worst possible scar outcome zero.

Scar pain parameters including pain dimensions, severity and interference were assessed. Pain dimensions were evaluated with the Neuropathic Pain Symptom Inventory (NPSI) (22), a validated inventory for evaluating the nature of pain, including spontaneous, paroxysmal and evoked pain and paraesthesia/dysesthesia. Pain dimension scores range from 0 to 10 with greater scores implying worse severity. Scar pain severity was assessed with the validated (23) Brief Pain Inventory severity scale (BPS) (24) and the Palmar Pain Severity Scale (PPS) (25). The BPS is calculated as the mean of present pain and the least, worst, and average pain over the last week, rated on an 11-point scale from 0 (no pain) to 10 (pain as bad as you can imagine). The PPS is a validated, patient-completed pain severity rating and was the primary outcome measure for identifying scar pain and correlation with objective scar measures. Scar pain interference was assessed with the Palmar Pain Interference Scale (PPI) (25), rated from none (zero) to extremely (100).

Psychological factors were evaluated using STarT Back Hands (**Supplementary Data 1**). This was a modification of the STarT Back Psychology Scale, a short psychological screening tool. The STarT Back Psychology Scale evaluates fear of movement, anxiety, catastrophizing and depression on a binary outcome, agree or disagree (26) and overall bothersomeness on a 5 point Likert scale, rated not at all to extremely. A score of four or five of five correlates with higher risk of poor functional outcome (27) and was interpreted as such in this study. The STarT Back tool was developed to screen primary care patients with low back pain for prognostic indicators relevant to providing personalized care. However, the measure was recently investigated in patients undergoing trapeziectomy surgery and identified a subgroup of patients at ‘high-risk’ who reported significantly worse outcomes(28). Given the growing evidence that higher levels of anxiety (29, 30) and catastrophic thinking in relation to pain (29, 31) are associated with poorer hand surgery outcome, validation of brief psychological screening tools is an important step in personalized medicine.

### Investigator-completed measures

Scar morphology (vascularity, pliability, pigmentation, height, relief and surface area) was evaluated with the Observer Scar Assessment Scale (OSAS) (32). Morphology dimensions are rated on a 10-point Likert scale, with 1 the same as normal skin and 10 the worst imaginable. The OSAS is validated for use in linear scars and has acceptable reliability (ICC for single parameters: 0.89–0.96) (11).

Infrared skin thermometry was used to evaluate peri-scar inflammation. Using an EXTECH Instruments dual laser InfraRed thermometer (model 42512), skin temperature was recorded three times. Mean temperature was reported for scar and a comparable site on the contralateral hand. Infrared skin thermometry is a reliable clinical assessment tool; an increase in skin temperature of 3° Fahrenheit is significant and suggests inflammation or infection (33).

Scar pliability was evaluated using a Checkline electromatic RX-1600-OO Type Durometer. Durometry is a reliable measure of the elastic and mechanical properties of scar and normal skin (34, 35). Indentional load is quantified with a retractable probe, determining tissue hardness.

Indentional load is dependent on viscoelastic properties and test duration. The durometer was applied manually, perpendicular to skin. Tissue firmness was expressed in arbitrary units from 0 to 100; with 100 equating to maximal firmness. Three trials were completed and mean calculated after one second (initial hardness) and 15 seconds (plasticity/creep) (36) and relative difference between the scar and contralateral hand determined.

### Quantitative Sensory Testing (QST)

Mechanical and thermal pain detection thresholds were evaluated with the German Research Network on Neuropathic Pain (DFNS) quantitative sensory testing (QST) protocol (37) by a trained examiner (DLK). This protocol is validated and widely used to assess somatosensory function (38). Participants were seated with the test hand supported on a table and introduced to procedures before testing. Assessment was first conducted on the contralateral limb in a comparable area. For all detection threshold measures, three trials were performed, and mean reported.

Mechanical pain threshold (MPT), (mechanical hyperalgesia) (**Fig 1A**) was evaluated using blunt, spring-loaded probes with forces ranging from 8 to 512 mN (pinprick stimulator, MRC, Heidelberg, Germany) and using a method of limits protocol; threshold is calculated as the geometric mean of five series of ascending and descending stimuli. Mechanical pain sensitivity (MPS) was evaluated using the same weighted probes. Participants rated pain from the range of stimulators on a scale of 0 for no pain and 100 for the worst pain imaginable. Dynamic mechanical allodynia (DMA) was assessed by participant pain ratings (0-100) to contact with a cotton wisp, a cotton bud (Q-Tip) and a standardised brush designed to produce minimum friction (Somedic, Sweden) (**Fig 1B**). Thermal pain thresholds were evaluated with a Somedic MSA thermal stimulator (Sweden) with an 18 mm^2^ metal Somedic thermode. This device has a baseline-temperature of 32°C, which increases or decreases in a pre-defined order (**Fig 1C**). Pressure pain threshold (PPT) was tested with a Wagner FDN 200 pressure algometer. The algometer was placed perpendicular to the skin, with a rubber tip against the participants’ scar. Pressure was gradually increased until the participant noted the change in sensation from pressure to discomfort, this value was recorded in Kg/cm^2^ (**Fig 1D**).

**Figure 1A-D.**
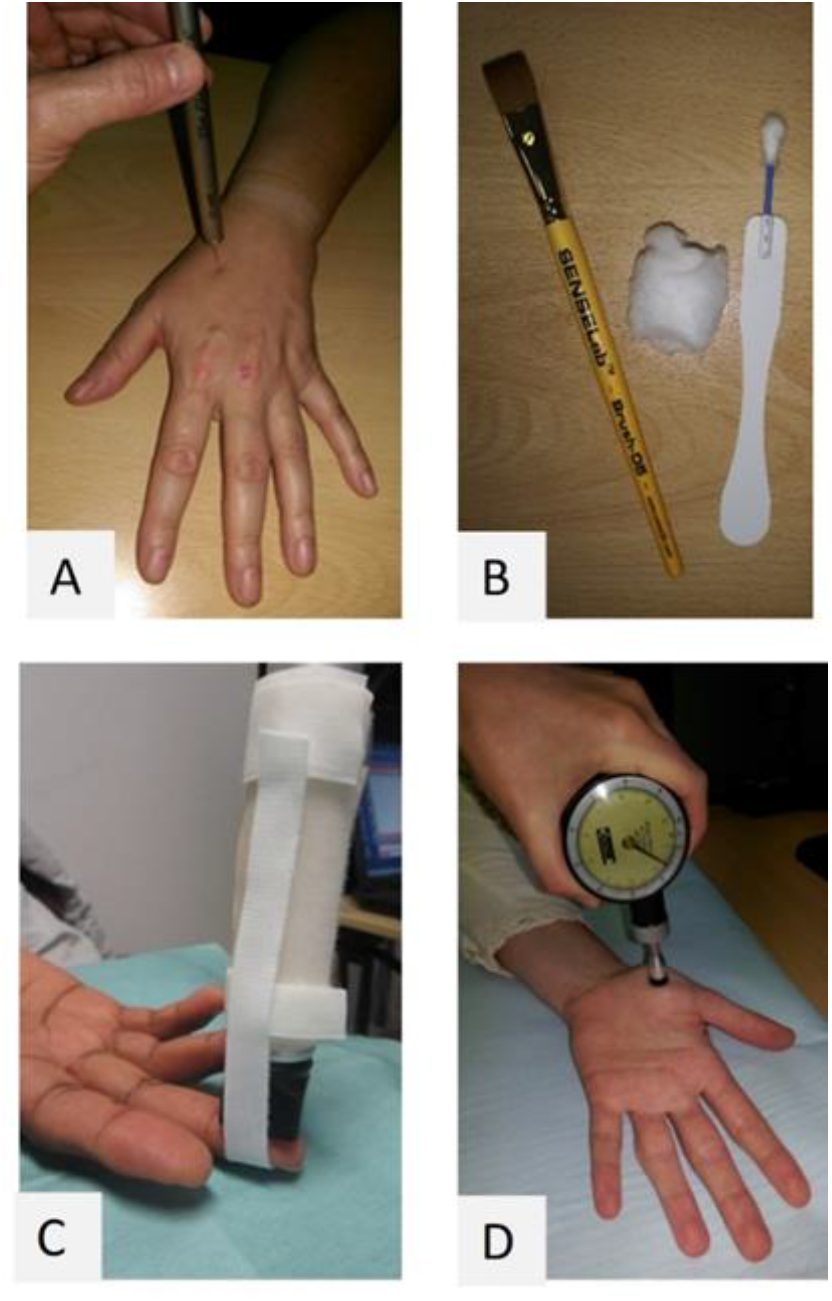
Quantitative Sensory Tests. A) Mechanical pain threshold, mechanical pain sensitivity; B) dynamic mechanical allodynia; C) cold and heat pain detection threshold; D) pressure pain threshold.

Statistical analysis was completed using IBM SPSS version 28. Participant characteristics, demographics and distribution of measures were summarized using descriptive statistics. Normality of data was assessed visually with histograms and statistically with the Kolmogorov-Smirnov test.

Continuous measures were reported as means (standard deviations) or medians (interquartile range); categorical data as counts (percentages). Change in measures was explored with paired– samples t-tests or Wilcoxon Signed Rank Test depending on data distribution. Differences between NPSI domains, symptoms and evocative stimuli scores were explored with the Friedman test. Effect size (Cohen’s d) and 95% confidence intervals (CI) are presented. Pearson or Spearman correlations were used to identify associations between the Palmar Pain Scale (primary outcome measure) and clinician-completed objective outcomes. Statistical significance was set at p< .05.

## RESULTS

The study opened February 25^th^ 2019 and was scheduled to close on December 31^st^ 2020. However, in keeping with national guidance, recruitment closed on April 1^st^ 2020 due to the COVID-19 pandemic. Prior to suspension, twenty-one participants (22% of eligible patients) provided informed consent and enrolled. Two participants (9%) dropped out after baseline assessment (one moved away; one became unwell). At study closure, 13 participants completed baseline and follow up assessments **(Figure 2)**.

**Fig. 2.**
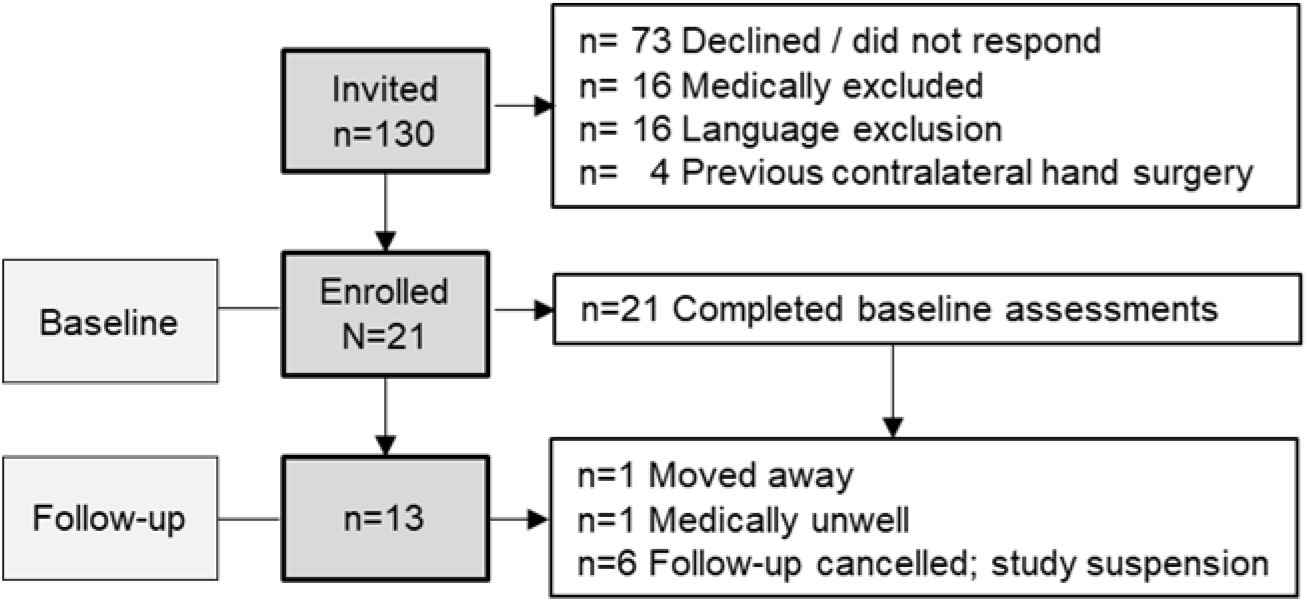
Study recruitment and enrolment

Demographic and health parameters are reported in **Table 1**. Prolene sutures were removed at mean (standard deviation) 13.4 (2.2) days. Baseline assessments were performed at mean (standard deviation) 32.6 (14.4) days; follow-up at 132 (18.5) days. At follow-up, there were no scar healing complications and no participants reported receiving clinical care for their scar. However, 9 participants (70% of participants followed up) reported performing scar care based on the advice of others or secondary to internet information (scar massage n=6; silicone gel n=2; Bio-Oil n=1).

**Table 1.** Key demographic and health parameters

|  |  |
| --- | --- |
| Age mean years (SD) | 60 (15.6) |
| Female sex n (%) | 16 (76) |
| **Surgical procedure n (%)** |  |
| Carpal tunnel decompression | 12 (57) |
| Trigger finger release | 3 (14) |
| Trapeziectomy | 2 (10) |
| Dorsal wrist ganglion | 1 (5) |
| Dupuytren's fasciectomy | 1 (5) |
| Interphalangeal joint cyst excision | 1 (5) |
| Trigger thumb release | 1 (5) |
| **Wound closure n (%)** |  |
| Dissolving suture | 3 (14) |
| Prolene | 18 (86) |
| **Surgical hand n (%)** |  |
| Dominant | 14 (67) |
| **Health parameters n (%)** |  |
| Diabetes | 5 (24) |
| Thyroid disease | 2 (10) |
| **Smoking** |  |
| never smoked | 13 (62) |
| current smoker | 0 (0) |
| **Employment status n (%)** |  |
| Employed | 6 (28) |
| Self-employed | 2 (10) |
| Unemployed | 3 (14) |
| Retired | 10 (48) |
| **Profession** |  |
| Elementary occupations | 1 (5) |
| Sales; customer service | 1 (5) |
| Administrative & secretarial | 3 (14) |
| Associate professional & technical | 2 (10) |
| Professional occupations | 12 (57) |
| Managers, directors | 2 (10) |
| SD, standard deviation |  |

### Participant-Reported Scar Pain Parameters

Scar pain parameters are reported in **Table 2**. Scores for the Palmar Pain Severity (PPS) indicate participants experienced, on average, mild scar pain at baseline and a statistically significant improvement to very mild pain at follow-up, demonstrating a large effect (d= 1.13; CI 0.41-1.82). Similarly, Palmar Pain Interference (PPI) was moderate at baseline and demonstrated a significant change with large effect (d= 0.86; CI 0.20-1.49) to minimal interference at follow up. The dispersion of scores for the Brief Pain Inventory (BPI) scales and mean score are illustrated in **Figure 3**.

**Table 2.**
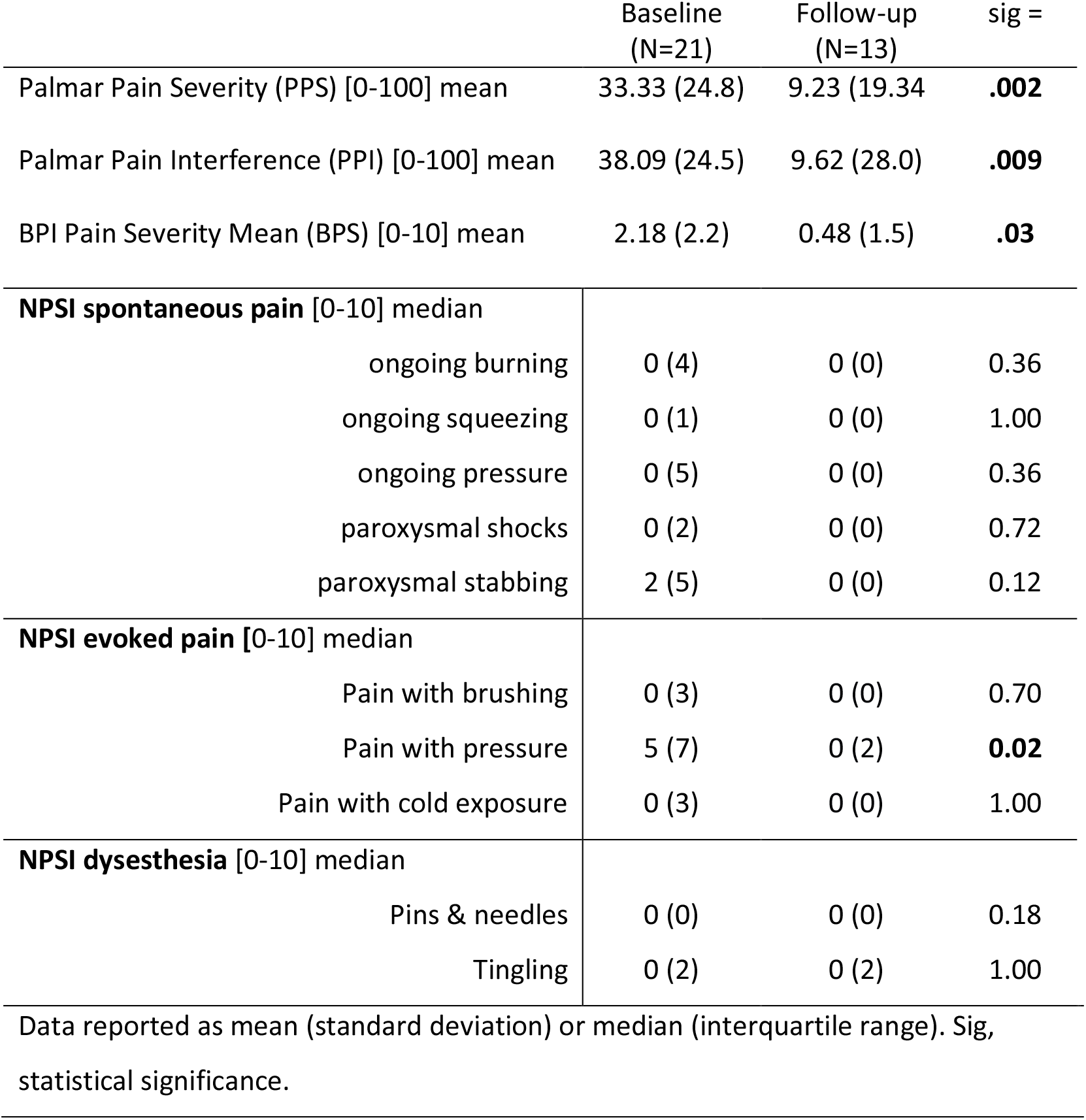
Patient-reported scar pain parameters at baseline and follow up

|  | Baseline<br>(N=21) | Follow-up<br>(N=13) | sig = |
| --- | --- | --- | --- |
| Palmar Pain Severity (PPS) [0-100] mean | 33.33 (24.8) | 9.23 (19.34) | **.002** |
| Palmar Pain Interference (PPI) [0-100] mean | 38.09 (24.5) | 9.62 (28.0) | **.009** |
| BPI Pain Severity Mean (BPS) [0-10] mean | 2.18 (2.2) | 0.48 (1.5) | **.03** |
| **NPSI spontaneous pain [0-10] median** |  |  |  |
| ongoing burning | 0 (4) | 0 (0) | 0.36 |
| ongoing squeezing | 0 (1) | 0 (0) | 1.00 |
| ongoing pressure | 0 (5) | 0 (0) | 0.36 |
| paroxysmal shocks | 0 (2) | 0 (0) | 0.72 |
| paroxysmal stabbing | 2 (5) | 0 (0) | 0.12 |
| **NPSI evoked pain [0-10] median** |  |  |  |
| Pain with brushing | 0 (3) | 0 (0) | 0.70 |
| Pain with pressure | 5 (7) | 0 (2) | **0.02** |
| Pain with cold exposure | 0 (3) | 0 (0) | 1.00 |
| **NPSI dysesthesia [0-10] median** |  |  |  |
| Pins & needles | 0 (0) | 0 (0) | 0.18 |
| Tingling | 0 (2) | 0 (2) | 1.00 |

Data reported as mean (standard deviation) or median (interquartile range). Sig, statistical significance.

**Fig 3.**
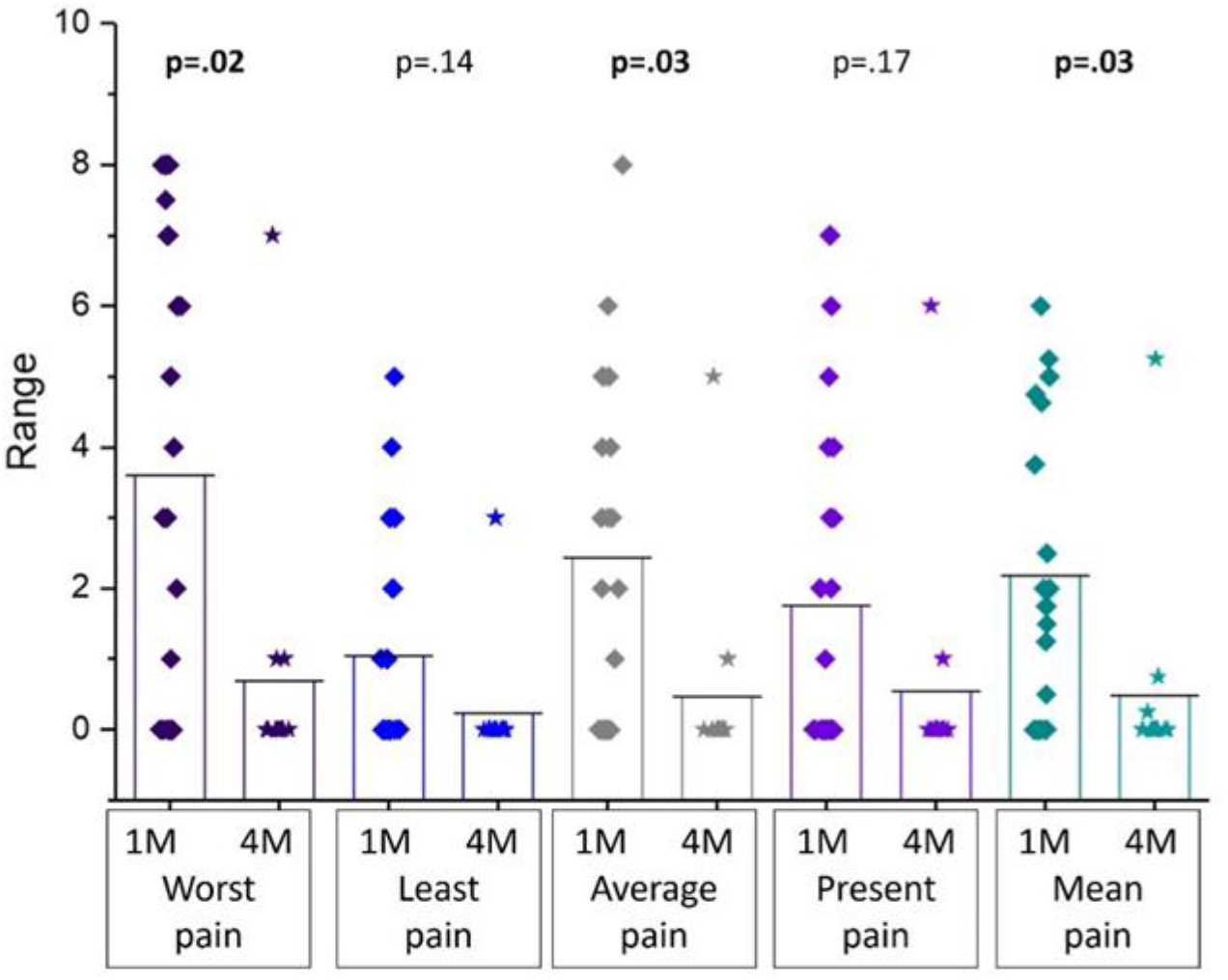
BPI Pain Severity Scale scores. Box represents mean (top line) and standard deviation, individual scores are represented by each dot. The mean of four scales (worst, least, average and present pain) is reported.

Change in scores for worst pain, average pain and mean pain rating were statistically significant with a moderate effect size (d=.75 [CI 0.17-1.35]; d=.69 [CI 0.07-1.29]; d=.68 [CI 0.07-1.27]), respectively. There was a large, significant association between the Palmar Pain Severity score and Brief Pain Inventory severity mean score (BPS) (*r*= .72, P<001).

Heterogeneity and change in pain symptoms were explored using the Neuropathic Pain Symptom Inventory (NPSI) (**Fig 4)**. At baseline, median (interquartile range) total NPSI score was 3 (8) and at follow-up diminished to 0 (3), however change was not significant (p=.31). Comparing symptoms of spontaneous pain; ongoing burning and pressure pain were more severe than squeezing pain (p=.04; p=.05), respectively. Comparing symptoms of spontaneous paroxysmal pain, stabbing pain was more severe than electrical pain (p=.02). For evoked pain, pressure evoked pain was greater than pain provoked by brushing or cold (p <.001). At baseline, 5 (24%) of participants reported pins & needles in the painful scar area; 9 (43%) reported tingling. Change scores for NPSI symptoms were statistically significant only for evoked pressure pain (p=.02).

**Fig 4.**
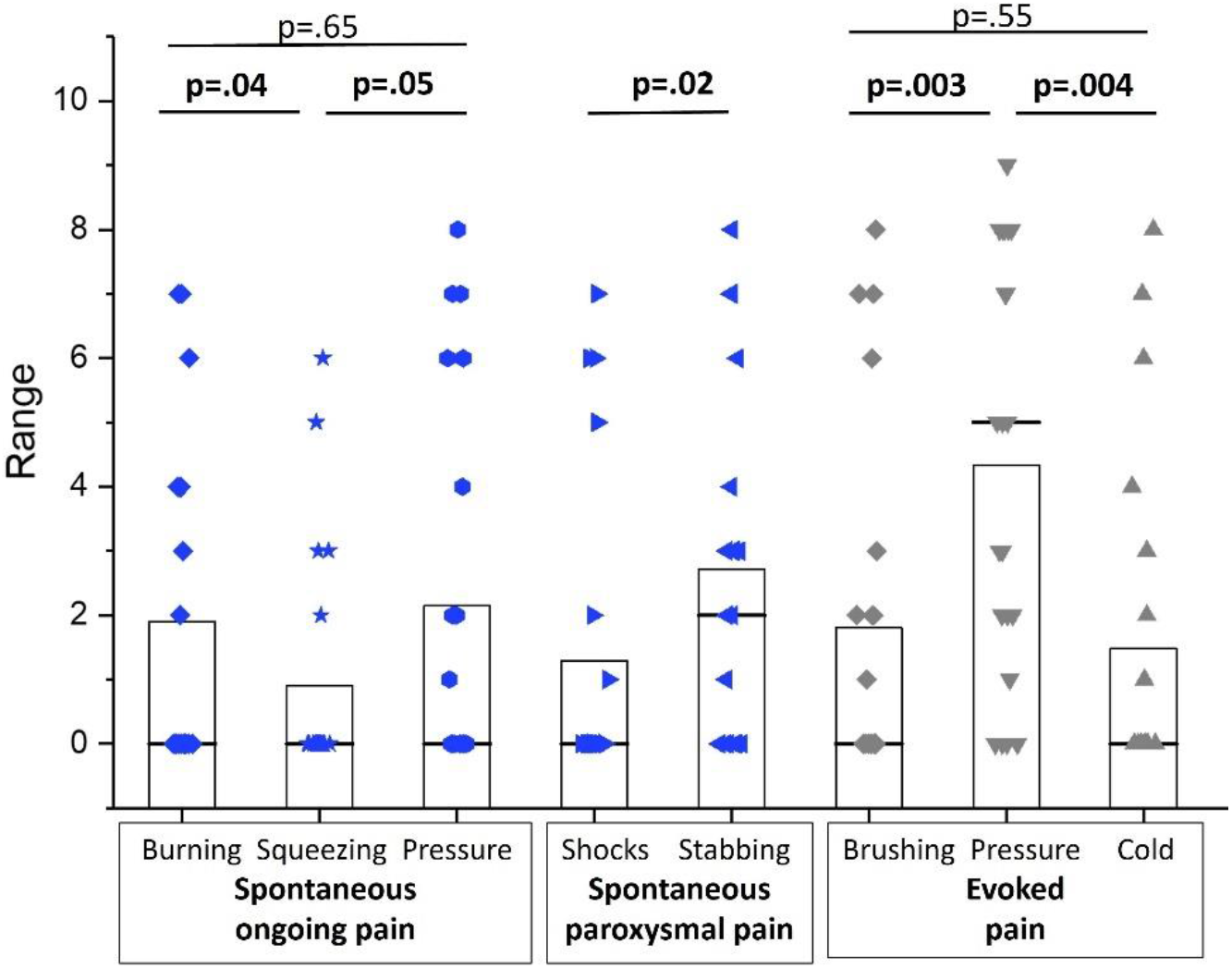
NPSI baseline scores. Box represents interquartile range, black horizontal line the median, individual scores are represented by each dot.

Participant rated scar appearance, consciousness and satisfaction with symptoms was evaluated with Patient Scar Assessment Questionnaire (PSAQ) subscales (**Fig 2**). Change scores for appearance were significant with a moderate effect (d=.73; CI 0.07-1.35), change in scar consciousness was significant with a large effect (d=1.09; CI 0.35-1.79). At 4 months post-surgery, participant global rating of scar outcome was median (interquartile range) 88.67 (13.66).

**Fig 2.**
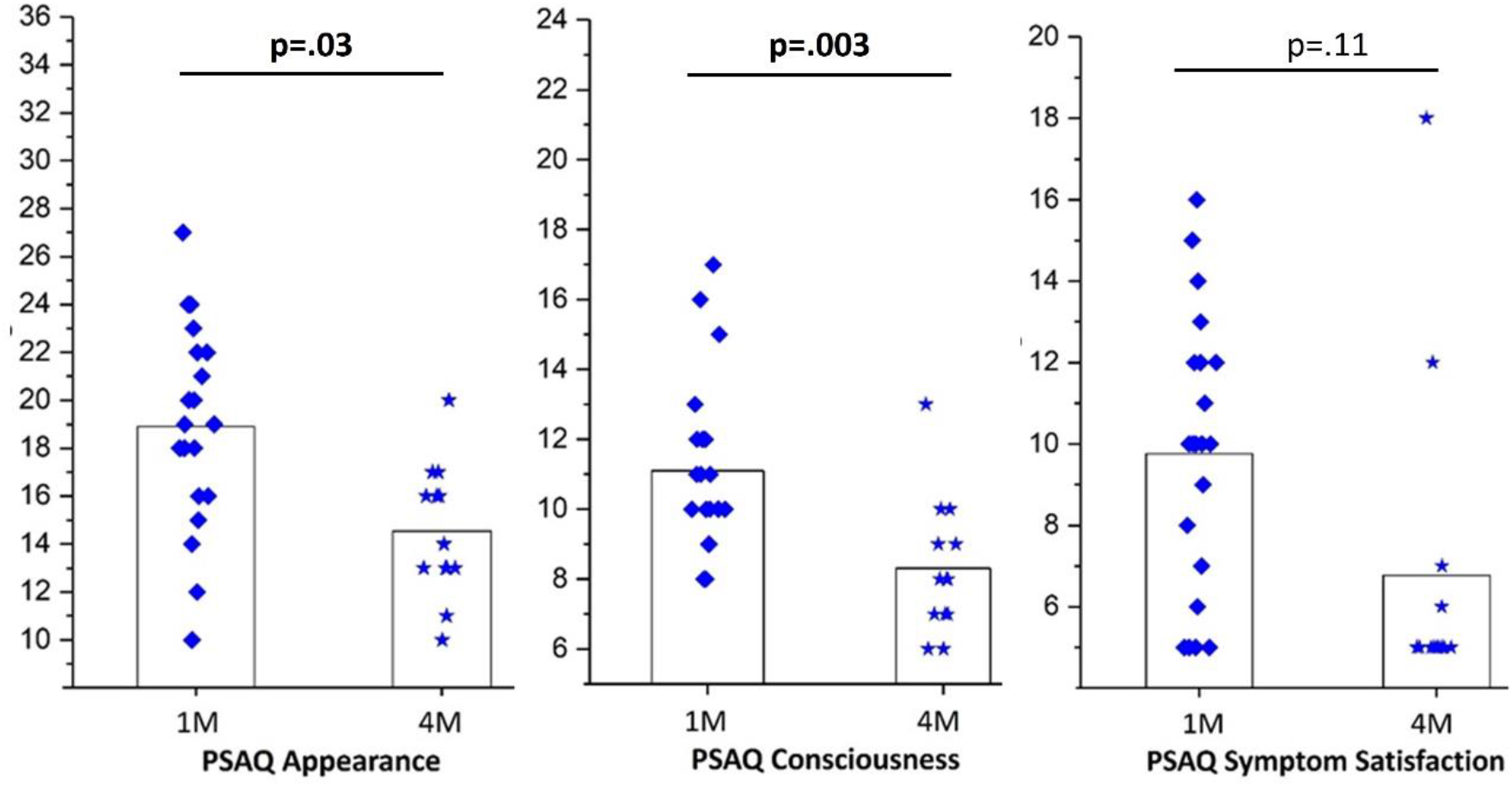
PSAQ subscales. Top line of the box represents the mean, the box the standard deviation and each dot a participant.

STarT Back Hands identified 2 (10%) participants at high risk of poor functional outcome, based on a score of ≥4 of a possible 5. Fourteen (67%) participants scored zero out of 5. Baseline StarT Back Hands scores were moderately correlated with participant reported scar pain r=.437, p=0.04, and strongly correlated with scar interference r = .708, p< .001.

### Investigator Completed Scar Evaluation

Scar morphology was evaluated with the Observer Scar Assessment Scale (OSAS). Baseline OSAS score mean (standard deviation) was 13.86 (4.18) and follow-up score 12.92 (2.1); change was not significant (p=.13). Scar morphology was not associated with patient-reported scar pain at baseline (p=.74) or follow-up (p=.8).

Scar inflammation was assessed with infrared thermometry. At baseline, median (IQR) scar temperature was 34.6°C (3.2), contralateral hand was 34.5°C (2.8), the difference was not significant (p=0.39). At follow-up, scar temperature was 33.2°C (2.0) and the contralateral hand was 33.03°C (1.8). The difference in temperature between assessments was not significant (p=0.78), nor was the difference between hands (p=0.60).

Scar pliability was evaluated with durometry. At baseline and follow-up, differences between unaffected skin and scar were statistically significant, demonstrating decreased pliability in the scar (**Table 3**). However, change in scar pliability from baseline to follow up was not significant (unaffected skin 1 p=0.51; unaffected skin 15 p=0.72; scar 1 p=0.54; scar 15 p=0.54). Importantly, neither baseline scar pliability measures (scar 1; scar 15) were associated with patient-reported scar pain (r =-0.138, p=0.55; r=-.138, p=0.47); respectively.

**Table 3.** Durometry assessment of scar pliability

| Table 3. Durometry assessment of scar pliability |  |  |  |
| --- | --- | --- | --- |
|  |  | mean (sd) | p= |
| Baseline<br>(N=21) | Skin 1<sup>a</sup> | 31.83 (6.85) | <.001 |
|  | Scar 1 | 44.65 (10.12) |  |
|  | Skin 15<sup>b</sup> | 31.49 (6.70) | <.001 |
|  | Scar 15 | 42.98 (9.62) |  |
| Follow-up<br>(N=13) | Skin 1 | 33.85 (7.86) | <.001 |
|  | Scar 1 | 47.82 (9.21) |  |
|  | Skin 15 | 33.05 (7.86) | <.001 |
|  | Scar 15 | 46.10 (7.85) |  |

‘Skin’ pertains to unaffected skin in the anatomically comparable region on the contralateral limb. a) Measurement ‘1’ is completed at one second; b) measurement ‘15’ is completed at 15 seconds.

QST was completed at baseline and follow-up (**Table 4**). Differences in Mechanical Pain Threshold (MPT) for scar compared to unaffected contralateral skin were not significant and MPT was not correlated with the PPS (*r* = -0.207; p=0.40). In contrast, the difference in Mechanical Pain Sensitivity (MPS) between hands was significant at baseline and the association of MPS with the PPS approached significance (*r* = 0.41; p=0.06). Dynamic Mechanical Allodynia (DMA), assessed at baseline and follow up, indicated no participant presented with DMA, i.e., no participant scored any of fifteen stimuli exposure as >0. Thermal pain detection and Pressure Pain Threshold (PPT) were assessed at follow up. The difference in cold pain threshold (CPT) between hands was significant, demonstrating cold hyperalgesia at the scar. CPT was not associated with patient reported scar pain (p=.34), however there was a significant positive association with patient-reported scar pain evoked by cold exposure on the NPSI questionnaire (*r* = .56; p = .05). Differences between sites for heat pain threshold (HPT) were not statistically significant and HPT was not associated with patient reported scar pain. PPT was tested in 7 of 13 (54%) participants at follow up. Per protocol(39), PPT is tested over soft tissue. Therefore, where scars were over a bony prominence, participants were excluded. PPT was median (IQR) 412 (75) kg/cm^2^ at the unaffected skin, 353 (294) kg/cm^2^ at the scar, demonstrating decreased PPT, or increased pressure pain sensitivity at the scar.

**Table 4.** Pinprick and thermal evoked pain measures

| Table 4. Pinprick and thermal evoked pain measures |  |  |  |
| --- | --- | --- | --- |
|  | Test site | Baseline | Follow-up |
| **Mechanical Pain Threshold (MPT)** | Skin | 157.6 (142.2) | 84.5 (223.34) |
|  | Scar | 128.0 (133.4) | 78.8 (85.69) |
|  | *sig* | P=0.21 | P=0.21 |
| **Mechanical Pain Sensitivity (MPS)** | Skin | 0.24 (0.17) | 0.32 (0.43) |
|  | Scar | 0.33 (0.39) | 0.29 (0.46) |
|  | *sig* | **P=0.04** | P=0.11 |
| **Cold Pain Threshold (°C) (CPT)** | Skin | NT | 7.17 (17.4) |
|  | Scar | NT | 24.07 (16.4) |
|  | *sig* | NT | **p=.05** |
| **Heat Pain Threshold (°C) (HPT)** | Skin | NT | 44.30 (8.6) |
|  | Scar | NT | 41.57 (3.7) |
|  | *sig* | NT | p=0.12 |
| 'Skin' pertains to unaffected skin in the anatomically comparable region on the contralateral limb. NT, not tested; Sig, statistical significance. Data reported with median (interquartile range). |  |  |  |

## Discussion

This study took a novel approach to investigating scar hyperaesthesia in adult patients following elective hand surgery for acquired conditions. This exploratory work aimed to identify objective clinical outcome measures associated with patient-reported scar pain and participant-reported measures that captured heterogeneity in scar symptoms for use in future studies.

Pain intensity was explored with the Palmar Pain Severity score (PPS) and Brief Pain Inventory severity score (BPS). Both scores were responsive to change over time and highly associated suggesting the PPS is a valid measure of scar pain intensity in other than carpal tunnel surgery populations. While the PPS uses one overall pain rating, in contrast the BPS includes four pain scales: the worst, least, average and present pain severity, and a mean pain rating. In this study, the baseline mean (standard deviation) BPS was 2.2 (2.2); in contrast the mean (standard deviation) for the worst pain scale was 3.6 (3.3). The distribution of baseline scores demonstrated that roughly 30% of participants rated their worst pain as ≥7 out of 10, highlighting severe pain that is undetected by mean or single rating pain intensity scales. Future scar trials may improve from using multiple scar pain severity ratings (worst, least, average and present pain) to capture heterogeneity in scar pain, and a composite score, as composite scores demonstrate greater reliability in pain research (40).

Scar pain interference was evaluated with the Palmar Pain Interference scale (PPI) which was responsive to change over time in a mixed elective hand surgery population. This is important, as the PPI is a clinically practical tool, being quick to administer and readily adaptable for face to face or virtual patient consultations. The scale may be implemented as a clinical scar outcome screening tool, identifying patients where further evaluation may be warranted.

We explored the nature and symptoms of post-surgical hand scar pain with the Neuropathic Pain Symptom Inventory (NPSI). NPSI total score diminished over time, however this change was not statistically significant. Exploring scar pain symptoms within the NPSI domains enabled the identification of important pain features in this sample. Burning and pressure were more severe symptoms of spontaneous pain than was squeezing. Stabbing spontaneous paroxysmal pain was more severe than shocks. Pressure evoked pain was more severe than brushing or cold evoked pain and had the highest pain rating across all symptoms. While there is scant evidence for the use of the NPSI for scar pain assessment, Huang, Wu (41) employed the NPSI in a study of the effectiveness of autologous fat grafting to alleviate neuropathic scar pain. The authors reported a statistically significant improvement in total NPSI scores, and similarly used the NPSI symptom scales to extrapolate dominant scar pain features. Use of the NPSI for the evaluation of scar pain symptoms in future studies may aide the interrogation of the drivers of persistent post-surgical scar pain and better inform treatment decisions for persistent scar pain.

In this sample, participant-rated scar appearance, consciousness and satisfaction with symptoms were evaluated with the relevant PSAQ subscales, demonstrating significant improvement in appearance and consciousness ratings over the three month follow up time. A large proportion of participants rated the appearance of their scar as poor and had a high degree of scar consciousness at baseline and follow up. This highlights the importance of patient-rated scar appearance; it is important that a patient-centred scar evaluation identify this psychological burden to ensure patients are adequately supported.

Scar morphology, as assessed with the OSAS, was not responsive to change and was not associated with participant reported scar pain. Scar pliability was significantly different to unaffected matched skin, however pliability measures did not change over time and likewise were not associated with participant reported scar pain. Scar inflammation, as assessed with infrared thermometry, identified no difference between scar and unaffected matched skin.

QST identified candidate psychophysical tests for quantifying scar thermal and mechanical sensitivity in future studies. Differences in Mechanical Pain Sensitivity (MPS), Cold Pain Threshold (CPT) and Pressure Pain Threshold (PPT) between scars and unaffected skin were identified and results were associated with patient reported scar pain, and patient reported cold evoked pain, respectively.

Quantification of thermal and mechanical pain sensitivity may support a personalised approach to scar pain treatment; change scores will support the evaluation of scar treatment effects in future studies. While no participant demonstrated Dynamic Mechanical Allodynia (DMA), this easily administered, and well tolerated test is useful for application in future studies.

There were no study protocol breaches or adverse events secondary to QST of post-surgical scars. PPT was not evaluated in participants where the surgical scar was over a bony prominence. Exploring sample demographics, the mean age was sixty years and participants were predominately retired or working in professional or managerial roles. There was little representation of those working in skilled trades, service workers or machine operators. In addition, protected characteristics were not included in the demographics so it is unknown if there is representation of the whole community in the sample. This highlights the need to engage a diverse patient steering group in study design to ensure study measures and methods are accessible to a representative sample of patients.

## Conclusion

Our findings in this cohort suggest patient reported scar pain is consistent with hyperaesthesia – an increased sensitivity to stimulation, including touch and thermal stimuli, with OR without pain. The PPS and PPI were responsive in a mixed sample of patients following planned hand surgery. Composite pain severity scores and the NPSI identified heterogeneity in pain severity and symptoms. The PSAQ was a responsive tool for quantifying patient rated scar appearance and consciousness. Psychophysical measures of MPS, CPT and PPT were well tolerated and identified mechanical and thermal pain sensitivity. While investigator evaluated scar morphology, pliability and inflammation were not associated with patient reported scar pain, the study sample size may not have been adequate to detect this difference.

## Data Availability

All data produced in the present work are contained in the manuscript.

---

**Supplementary data 1.**
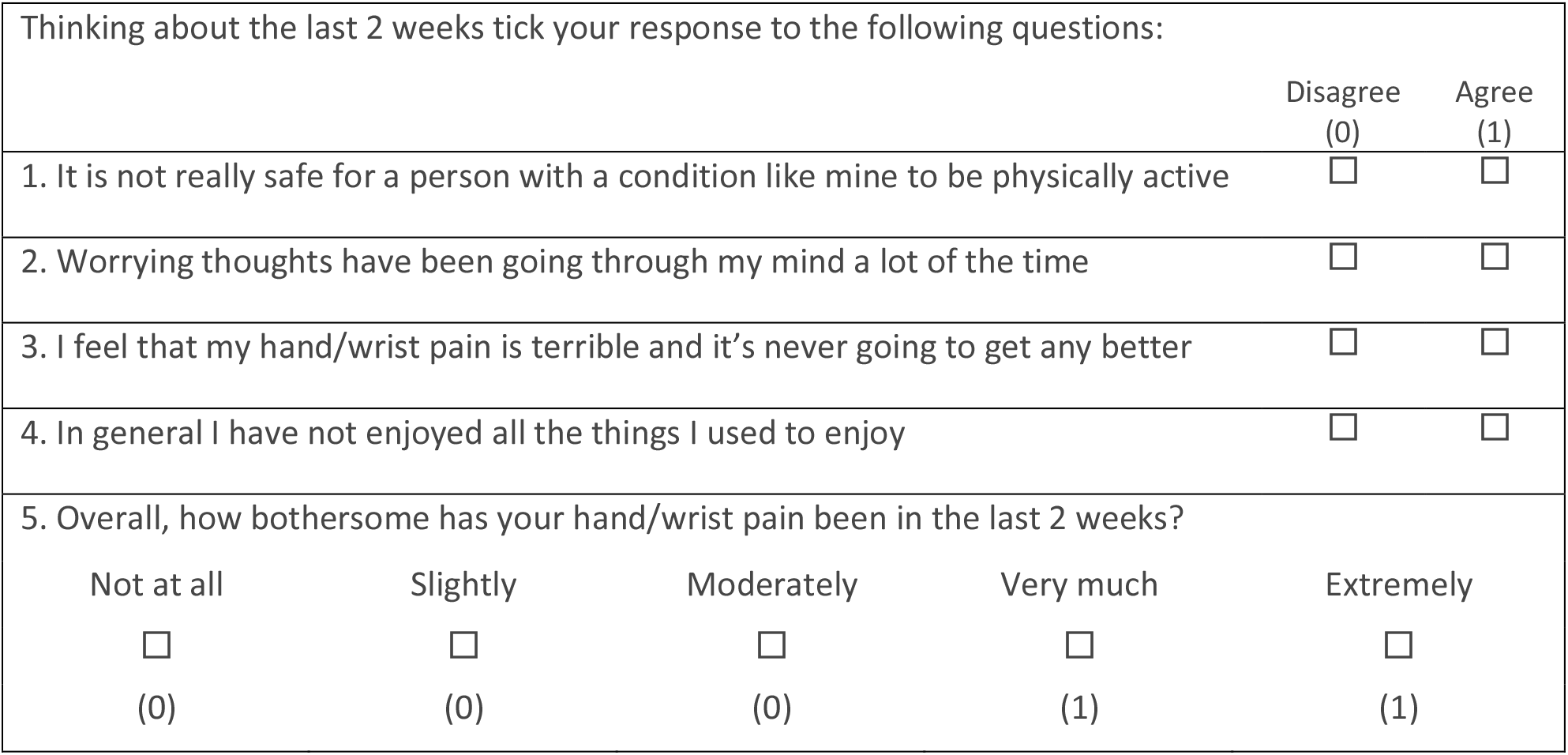
STarT Back Hands Questionnaire.

## Declarations

### 3. Informed consent

Written informed consent was obtained from study participants for their anonymized information to be published in this manuscript.

### 4. Ethical approval

HRA and Health and Care Research Wales approved this study (REC number: 18/LO/2161) on 20^th^ December 2018. This study was conducted according to the World Medical Association Declaration of Helsinki.

### 5. Data availability

Data supporting this study are included within the manuscript.

### 6. Contributorship

DLK & SH researched literature and conceived the study. DLK and CMA contributed to protocol development, gaining ethical approval and data analysis. DLK was responsible for patient recruitment and data acquisition. All authors reviewed and edited the manuscript and approved the final version of the manuscript.

## 7. Acknowledgements

We would like to thank patient collaborators Ms Maria Braga and Ms Veronica Green for their contribution to this research.

